# Serum ferritin and clinical outcomes in children undergoing pediatric cardiac surgery

**DOI:** 10.64898/2026.01.26.26344857

**Authors:** Naycka Onofre Witt Batista, Humberto Holmer Fiori, Nadia Camilato Ferraz Knop

## Abstract

**Introduction:** Hyperferritinemia is a prognostic marker in critical illness, but its role in postoperative outcomes of pediatric congenital heart defects remains poorly defined, especially in resource-limited settings. This study evaluated early serum ferritin as a predictor of outcomes after congenital heart surgery and its association with the PIM 3 score.

**Methods:** A single-center, prospective cohort study was conducted from April 2023 to October 2024 at a tertiary referral center in southeastern Brazil. Patients aged 29 days to 18 years, of both sexes, admitted to the PICU after congenital heart surgery were included and categorized as cyanotic or acyanotic. Statistical significance was defined as two-sided *p* < 0.05.

**Results:** A total of 105 patients were included. Median ferritin was higher in patients with PICU stays < 7 days (183 ng/mL; *p* = 0.004) and was significantly associated with a PIM 3 score ≥ 5% (642 ng/mL; *p* < 0.006). Cyanotic patients had longer PICU stays (11.0 vs. 7.2 days; *p* = 0.02), longer use of vasoactive drugs (3.8 vs. 2.6 days; *p* = 0.01), and accounted for all deaths (*p* < 0.001). Hemoglobin and hematocrit were also significantly higher in cyanotic patients (14 vs. 13 g/dL and 40% vs. 37%; *p* < 0.001).

**Conclusions:** Serum ferritin may serve as a marker of secondary outcomes and aid early risk stratification in congenital heart defects patients in the PICU.

## Introduction

A critically ill child presents with severe dysfunction of one or more organ systems, which, if not promptly and effectively treated, increases the risk of sequelae and death ^[1]^. Among the leading causes of mortality in the pediatric intensive care unit (PICU) is congenital heart defects, which, when compounded by infectious complications and heart failure, further heightens the risk of death in this patient population ^[2]^.

Congenital heart defects constitute a group of congenital abnormalities with a high birth prevalence, occurring in approximately 8 to 10 per 1.000 live births, and are associated with substantial functional impairment in affected patients ^[3]^. According to the Brazilian Ministry of Health ^[4]^, congenital heart defects are the second most common congenital anomaly in the country and represent a severe clinical condition that warrants further research to improve healthcare strategies.

Lorch et al. emphasized the importance of accounting for the clinical complexity of patients across different hospital settings and countries, as multiple factors influence disease incidence and clinical outcomes. The analysis of distinct populations admitted to the PICU and the identification of predictors of mortality and other adverse outcomes are essential for refining risk stratification and guiding clinical management.

Acute-phase biomarkers are widely used in pediatric risk scores, particularly C-reactive protein (CRP) and serum ferritin, both of which are associated with the inflammatory process and the monitoring of the therapeutic response ^[6]^. Ferritin, in particular, has demonstrated consistent prognostic value in children with sepsis, reinforcing its potential as a tool for early diagnosis and for guiding individualized treatment adjustments across different profiles of critically ill patients ^[7]^.

Despite its clinical relevance, few studies have systematically investigated the role of ferritin as an inflammatory biomarker in critically ill children following congenital heart surgery in the PICU. This study evaluated serum ferritin as an early predictor of clinical outcomes in children undergoing congenital heart surgery and examined its association with the PIM 3 score.

## Methods

### Study design and participants

This prospective cohort study was conducted at the Alzir Bernardino Alves Children’s and Maternity Hospital (HIMABA), a referral center for congenital heart surgery located in southeastern Brazil, between April 2023 and October 2024. Patients aged 29 days to 18 years, of both sexes, who were admitted to the PICU in the early postoperative period following congenital heart surgery were included. All procedures were performed by the same specialized surgical team, and each patient was included only once; in cases involving two index surgeries during the same hospitalization, only the first procedure was considered for analysis.

### Inclusion and exclusion criteria

Inclusion criteria required the measurement of postoperative serum ferritin and serum iron, as well as signed informed consent from the patient’s legal guardian. Exclusion criteria included noninvasive cardiac procedures, such as percutaneous interventions, recent blood transfusion (within four months prior to admission), chronic renal or hepatic disease, autoimmune disorders, hemophagocytic syndrome, or continuous use of immunosuppressive drugs, as these conditions could interfere with serum ferritin levels.

### Study protocol

Data collection was conducted prospectively over a 14-month period using an instrument specifically developed for this study. The collected variables included demographics (age in months, sex), anthropometric data (weight, in kilograms), clinical characteristics (type of congenital heart defect—cyanotic or acyanotic; need for and duration of mechanical ventilation; use and duration of vasoactive drugs; PICU length of stay, in days), and intraoperative factors (duration of cardiopulmonary bypass, in minutes). Clinical information was obtained from electronic and paper medical records from admission to discharge or death. Serum ferritin, serum iron, hemoglobin, and hematocrit levels were measured within 18 hours after surgery. Clinical severity and surgical risk were assessed using the Pediatric Index of Mortality 3 (PIM 3) and the Risk Adjustment for Congenital Heart Surgery 1 (RACHS-1) score, respectively.

### Risk scores

The Pediatric Index of Mortality 3 (PIM 3) is a prognostic model that estimates mortality risk in children admitted to the PICU based on initial clinical and laboratory parameters ^[8]^. In this study, PIM 3 scores were calculated using data extracted from hospital electronic systems and entered into a protocol-specific spreadsheet. The Risk Adjustment for Congenital Heart Surgery 1 (RACHS-1) score classifies surgical procedures into six risk categories according to expected mortality ^[9]^. In this study, patients were categorized according to the procedure performed, allowing for the analysis of surgical risk within the sample.

### Clinical outcomes and mortality

The clinical outcomes analyzed included PICU length of stay, duration of invasive mechanical ventilation, and duration of vasoactive drug use, all expressed in days and measured from admission following cardiac surgery until discharge or death. In-hospital mortality was considered the final outcome.

### Statistical analysis

Statistical analyses were performed using Epi Info, version 7.2.6.0. In the descriptive analysis, categorical variables were expressed as absolute and relative frequencies (%). Quantitative variables with normal distribution were presented as mean ± standard deviation (SD), whereas those with non-normal distribution were reported as median and interquartile range (IQR). For comparisons, the chi-square test or Fisher’s exact test (when appropriate) was applied for categorical variables; Student’s *t-test* (for two groups) or ANOVA (for more than two groups) for continuous variables with normal distribution; and the Kruskal–Wallis test for nonparametric data. Serum ferritin percentiles were analyzed in relation to the main PICU clinical outcomes—length of stay, duration of mechanical ventilation, and duration of vasoactive drug use (all expressed in days) —as well as mortality and the PIM 3 prognostic score. A *p*-value ≤ 0.05 was considered statistically significant.

### Ethical considerations

The study was conducted in accordance with the ethical principles established by Resolution No. 466/2012 of the Brazilian National Health Council. It was approved by the Research Ethics Committee of the Pontifical Catholic University of Rio Grande do Sul (PUCRS) under protocol number 67563123.5.0000.5336, on March 8, 2023. Informed consent was obtained from all participants included in the study.

## Results

A total of 105 patients with congenital heart defects were included, ranging in age from 29 days to under 18 years. Participants were followed until death or discharge from the unit, with the possibility of transfer to an intermediate care unit when indicated. All deaths occurred in the PICU, where withdrawal of life support is not a common practice.

Table 1 shows the comparison of demographic, anthropometric, laboratory characteristics and clinical outcomes of patients with congenital heart defects, as well as the analysis between cyanotic and acyanotic subgroups. No statistically significant differences were found in demographic or anthropometric variables between the subgroups. Regarding clinical outcomes, cyanotic patients had longer PICU stays (11.0 vs. 7.2 days; *p* = 0.02) and longer durations of vasoactive drug use (3.8 vs. 2.6 days; *p* = 0.01) compared with acyanotic patients. All observed deaths occurred in the cyanotic group (*p* < 0.001).

**Table 1.** Demographic, Anthropometric, Laboratory Characteristics, and Clinical Outcomes of Cardiac Patients Admitted to the PICU.

| Characteristics | Cardiac<br>(n = 105) | Cyanotic<br>(n = 20) | Acyanotic<br>(n = 85) | *p*-value |
| --- | --- | --- | --- | --- |
| **Demographic** |  |  |  |  |
| Age, months – mean (SD) | 46 (53) | 34 (45) | 48 (55) | 0,28 |
| Male sex, *n* (%) | 46 (43,8%) | 11 (55%) | 35 (41,2%) | 0,32 |
| **Anthropometric** |  |  |  |  |
| Weight, kg – mean (SD) | 16,1 (14,9) | 14,3 (13,4) | 16,6 (15,3) | 0,53 |
| **Clinical outcomes** |  |  |  |  |
| MV, *d* – mean (SD) | 1,5 (1,7) | 2,3 (3,0) | 1,3 (1,2) | 0,15 |
| VD, *d* – mean (SD) | 2,8 (1,6) | 3,8 (2,0) | 2,6 (1,5) | **0,01\*** |
| PICU LOS, *d* – mean (SD) | 7,9 (4,4) | 11,0 (7,5) | 7,2 (2,8) | **0,02\*** |
| Death, *n* (%) | 4 (3,8%) | 4 (20%) | 0 (0%) | **< 0,001\*** |
| **Laboratory** |  |  |  |  |
| Serum iron, µg/dL – median (IQR) | 50 (28-104) | 47,5 (22–56) | 52 (29–109) | 0,43 |
| Serum ferritin, ng/mL – median (IQR) | 158 (91-262) | 140 (109–217) | 159 (90–293) | 0,87 |

**Notes:** % = percentage; *d* = days; IQR = interquartile range; kg = kilograms; LOS = length of stay; MV = mechanical ventilation; *n* = number of patients; ng/mL = nanograms per milliliter; PICU = pediatric intensive care unit; SD = standard deviation; µg/dL = micrograms per deciliter; VD = vasoactive drugs.
\* *p*-values < 0.05 were considered statistically significant.
**Source:** Study data.

### Laboratory findings

Regarding laboratory characteristics, median serum iron and serum ferritin were similar between the subgroups (Table 1). In the analysis, median hemoglobin and hematocrit values were significantly higher in cyanotic than in acyanotic patients (14 vs. 13 g/dL, *p* < 0.001; and 40% vs. 37%, *p* < 0.001, respectively) (Table 2). Conversely, red blood cell indices (MCV, MCH, RDW), which are indicators of microcytic and hypochromic anemia, showed no significant differences between the subgroups.

**Table 2.** Red Blood Cell Parameters and Hematimetric Indices According to Cardiac Patient Groups in the Study.

| Index | Median (IQR) |  | *p*-value |
| --- | --- | --- | --- |
|  | Acyanotic | Cyanotic |  |
| Hematocrit (%) | 37 (33-40) | 40 (37-44) | < 0,001* |
| Hemoglobin (g/dL) | 13 (11,4-13,6) | 14 (13,2-15,2) | < 0,001* |
| MCV (fL) | 81 (78-85) | 80 (77-84) | 0,89 |
| MCH (pg) | 28 (26-29) | 28 (26-29) | 0,04 |
| RDW (%) | 15 (14-16) | 15 (14-16) | 0,83 |

**Notes:** % = percentage; fL = femtoliters; g/dL = grams per deciliter; MCH = mean corpuscular hemoglobin; MCV = mean corpuscular volume; pg = picograms; RDW = red cell distribution width.
\* Values expressed as median (interquartile range, P25–P75).
\* *p*-values < 0.05 were considered statistically significant.
**Source:** Study data.

### RACHS-1 score and cardiopulmonary bypass time

Table 3 presents the distribution of RACHS-1 categories, with a predominance of surgical risk category 2 (47%), followed by categories 1 (28%) and 3 (25%). Median cardiopulmonary bypass (CPB) time progressively increased across higher risk categories, reaching its highest value in category 3 (Figure 1).

**Table 3.** Risk Categories of Surgical Cardiac Patients According to the RACHS-1 Score.

| Risk Category | Number of Patients (n = 105) | % of Total Surgical Patients |
| --- | --- | --- |
| 1 | 29 | 28% |
| 2 | 49 | 47% |
| 3 | 27 | 25% |
| 4 | 0 | 0% |
| 5 | 0 | 0% |
| 6 | 0 | 0% |

**Note:**
Classification adapted from Jenkins *et al.* (2002), based on surgical complexity of congenital heart disease.
**Source:** Study data.

**Figure 1.**
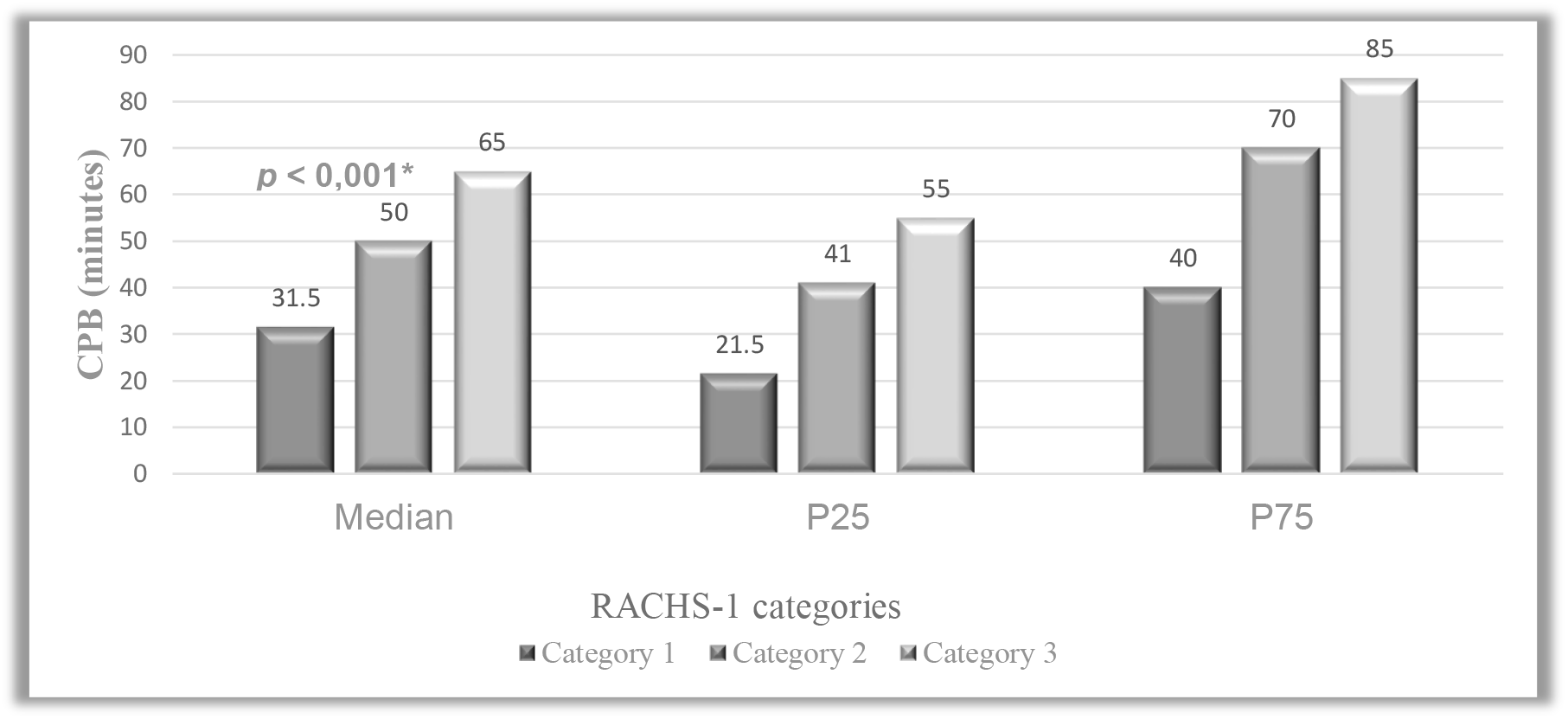
Cardiopulmonary bypass time according to RACHS-1 risk categories. **Note:** Values expressed as median (IQR). *Kruskal–Wallis, *p* < 0.001. **Source:** Study data.

**Figure 1.**
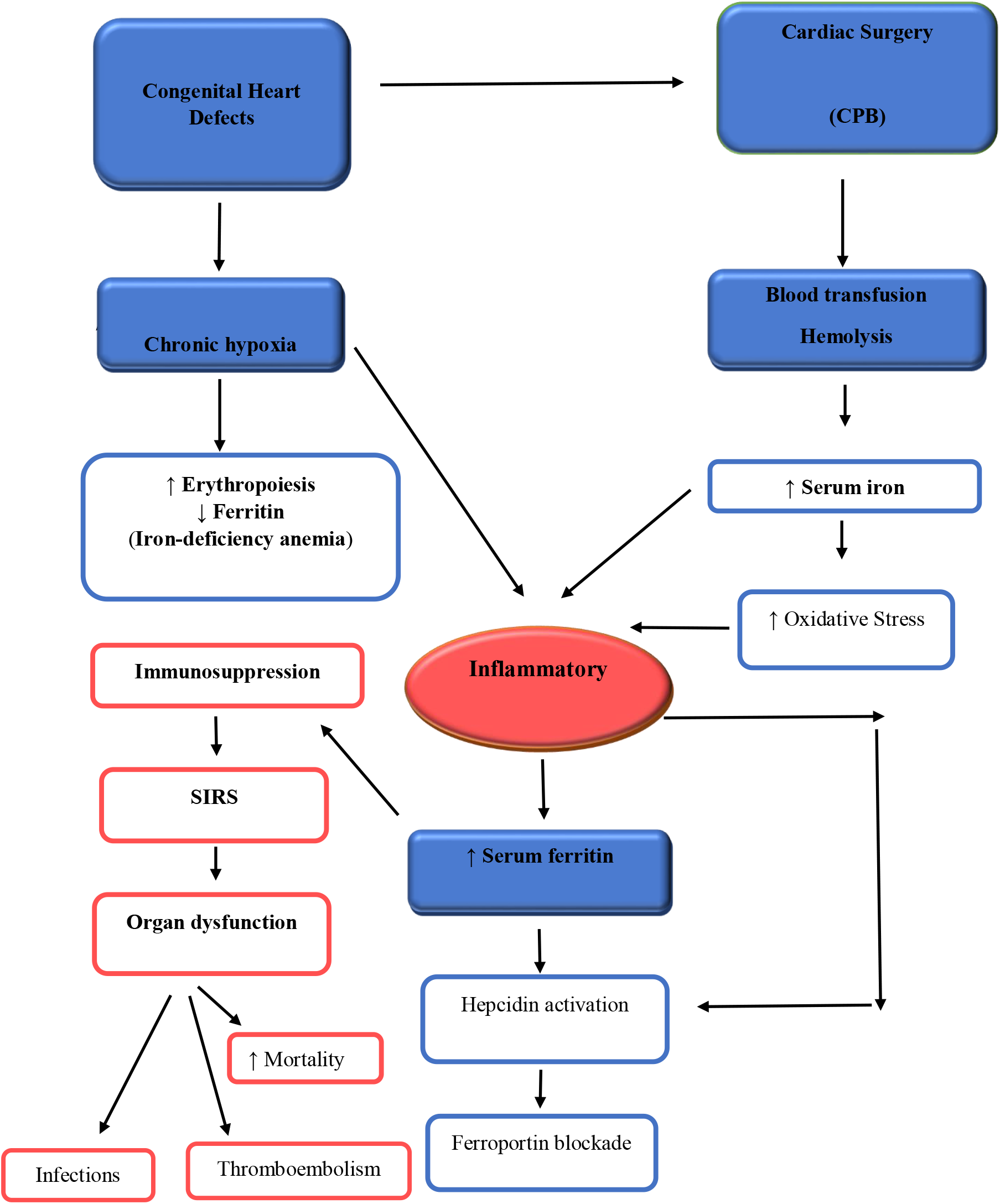
Schematic representation of iron and serum ferritin metabolism in patients with congenital heart defects undergoing cardiac surgery. **Note: CPB =** Cardiopulmonary bypass; SIRS = Systemic Inflammatory Response Syndrome. **Source:** prepared by the author (2025).

### Serum ferritin, clinical outcomes, and PIM 3

Regarding clinical outcomes, a statistically significant difference in median serum ferritin levels was observed in patients with PICU stays shorter than seven days (183 ng/mL; *p* = 0.004). In the analysis of the association between PIM 3 ≥ 5% and serum ferritin percentiles, a significant difference was also identified (642 ng/mL; *p* < 0.006). Conversely, no significant association was found between median postoperative serum ferritin levels and the outcomes of death, duration of vasoactive drug use, or length of mechanical ventilation, as shown in Table 4.

**Table 4.** Comparison of clinical outcomes and PIM 3 score according to serum ferritin percentiles in patients with congenital heart disease.

| Outcome | Median ferritin (ng/mL) | p25 | p75 | p-value |
| --- | --- | --- | --- | --- |
| **Death** |  |  |  | 0,65 |
| No (n=101) | 158 | 91 | 262 |  |
| Yes (n=4) | 174 | 116 | 422 |  |
| **PICU LOS** |  |  |  | **0,004\*** |
| < 7d (n=69) | 183 | 119 | 293 |  |
| ≥ 7d (n=36) | 107 | 67 | 210 |  |
| **MV** |  |  |  | 0,33 |
| < 7d (n=96) | 161 | 99 | 264 |  |
| ≥ 7d (n=9) | 76 | 63 | 183 |  |
| **VD** |  |  |  | 0,82 |
| < 3d (n=79) | 147 | 90 | 241 |  |
| ≥ 3d (n=26) | 195 | 114 | 333 |  |
| **PIM 3** |  |  |  | **0,006\*** |
| < 5% (n=102) | 157 | 90 | 246 |  |
| ≥ 5% (n=3) | 642 | 130 | 910 |  |

**Note:** % = percentage; d = days; LOS = length of stay; MV = mechanical ventilation; n = number of patients; ng/mL = nanogram per milliliter; PICU = pediatric intensive care unit; PIM 3 = *Pediatric Index of Mortality 3*; VD = vasoactive drugs.
\*Values of $p < 0.05$ were considered statistically significant.
**Source:** Study data.

Figure 2 integrates the main pathophysiological mechanisms related to congenital heart defects, iron metabolism, and cardiopulmonary bypass–associated surgical trauma that may alter postoperative serum iron and ferritin levels, including chronic hypoxia, hemolysis, oxidative stress, and systemic inflammatory response, all of which relate to the clinical outcomes observed.

## Discussion

In the present study, higher serum ferritin levels were associated with cardiac surgical patients who remained in the PICU for fewer than seven days, as well as with those presenting higher PIM 3 scores. Comparative analysis of the main clinical outcomes showed that cyanotic patients required longer use of vasoactive drugs, had longer PICU stays, and exhibited higher mortality rates compared with acyanotic patients. However, regarding mortality, serum ferritin did not demonstrate value as an early prognostic biomarker.

These findings are relevant, because, although other studies have analyzed serum ferritin levels in the context of sepsis and other conditions, few have specifically investigated its value as an early biomarker in patients with congenital heart defects, particularly in referral centers for pediatric cardiac surgery. In this context, serum ferritin levels may be directly associated with the acute postoperative inflammatory response and reflect the clinical severity of patients requiring greater hemodynamic support in the PICU.

Exposure to cardiopulmonary bypass (CPB) and blood transfusion during cardiac surgery induces hemolysis and alters iron metabolism in patients with congenital heart defects (figure 2), promoting an inflammatory state associated with poorer postoperative outcomes ^[10]^. Conversely, iron deficiency anemia, often underdiagnosed in these patients, stimulates erythropoiesis and depletes ferritin stores ^[11]^. These apparently opposing mechanisms coexist in surgical patients and may balance differently depending on the clinical context. In developing countries such as Brazil, the high prevalence of iron deficiency anemia may attenuate hyperferritinemia in this critically ill population, potentially explaining the more modest ferritin levels observed in this study despite the expected inflammatory response.

Guo et al. evaluated 457 children under three years of age who underwent cardiac surgery with CPB. On the first postoperative day, 10.9% had ferritin levels ≥ 250 ng/mL and 1.1% had levels ≥ 1.000 ng/mL. These findings indicate that early hyperferritinemia is relatively common after cardiac surgery with CPB and represents an important predictor of in-hospital complications, such as prolonged mechanical ventilation and extended length of stay.

PICU length of stay following cardiac surgery is widely described in the literature as a relevant prognostic marker. Roodpeyma et al. reported a mean stay of 4.9 days, with shorter stays associated with early mortality, particularly among infants and neonates with cyanotic congenital heart defects. In contrast, Belo et al. reported a mean stay of 16 days, showing a positive correlation between the number of cardiac malformations and length of stay. Although distinct, these findings highlight the influence of age and anatomical complexity on prognosis. In the present study, these factors were also reflected in the greater clinical severity of cyanotic patients, particularly those with higher ferritin levels, which were associated with PICU stays shorter than seven days, possibly resulting from early postoperative complications leading to higher mortality in this subgroup.

These results reinforce the association between elevated ferritin levels, systemic inflammation, and unfavorable outcomes, particularly in more complex congenital heart defects. However, in this study, ferritin concentrations above 3.000 ng/mL, previously associated with higher mortality in critically ill patients, were not observed ^[15]^. This finding suggests a less intense inflammatory response in the study population, possibly related to the predominance of elective acyanotic surgeries and shorter CPB durations performed by a specialized surgical team.

Patients undergoing more complex surgical procedures exhibit higher mortality rates, particularly when associated with cardiogenic shock (61.8%) or septic shock (35.5%) ^[16]^. Postoperative infection is especially relevant in younger children and in those with PICU stays shorter than six days, suggesting immunological vulnerability and rapid clinical deterioration ^[17,18]^.

International studies report significant mortality rates: 9.8% among neonates in Malaysia ^[19]^ and up to 12.2% in China, reaching 8.6% in cases of complex congenital heart defects ^[20]^. The findings of the present study corroborate these data, as all deaths occurred in cyanotic patients, most of whom were infants. Although specific causes were not assessed, this age group is widely recognized as being more vulnerable to postoperative hemodynamic and infectious complications. Survival of fewer than seven days, associated with higher serum ferritin levels, underscores the early clinical severity and intensity of the inflammatory response in the postoperative period. Compared with acyanotic patients, those with cyanotic congenital heart defects also had longer PICU stays, prolonged use of vasoactive drugs, and higher mortality rates, confirming that the combination of surgical complexity and younger age contributes to poorer outcomes.

The results of this study demonstrated that higher ferritin levels were associated with PIM 3

≥ 5%, indicating its potential as a complementary biomarker for the prognostic assessment of critically ill patients with congenital heart disease. Given the limitations of PIM 3 in subgroups of greater complexity, the incorporation of ferritin may enhance risk stratification, particularly in resource-limited settings, where clinical and surgical heterogeneity compromises the accuracy of traditional scoring systems.

Multicenter studies have demonstrated satisfactory performance of PIM 3 across various pediatric settings. In Italy, the score demonstrated good accuracy in all evaluated categories, including postoperative cardiac surgery patients ^[21]^. Similar findings were reported in a prospective study conducted in nine South African PICUs, which included a large number of surgical patients and showed adequate discriminatory capacity for predicting survival and death ^[22]^. In Argentina, although PIM 3 exhibited excellent discriminatory ability, mortality was underestimated, particularly among adolescents and those with complex chronic conditions ^[23]^.

Although useful, PIM 3 does not account for surgical complexity, which is addressed by the RACHS-1 score. In this study, category 2 predominated (47%), followed by category 1 (28%), a pattern similar to that reported by Nina et al., whereas Pelissari et al. observed a higher frequency of category 3, which was associated with a greater number of postoperative complications.

Several limitations must be acknowledged. The low mortality rate limited the analysis of death as a primary outcome, and the single-center design compromises external validity due to potential selection bias. Furthermore, the heterogeneity of congenital heart defects and the exclusion of neonates, a group particularly vulnerable to complex surgeries and higher mortality, may have restricted the generalizability of the results and contributed to an underestimation of the observed ferritin levels.

In the present study, no marked early hyperferritinemia or significant reduction in serum iron levels after cardiac surgery with cardiopulmonary bypass (CPB) was observed, suggesting a distinct pattern of iron metabolism, particularly in acyanotic congenital heart defects. Although no significant associations with the main clinical outcomes in the early postoperative period in the PICU were identified, an association was observed between elevated serum ferritin levels and PIM 3 scores ≥ 5%. Future investigations, especially in more complex subgroups such as cyanotic patients, will be essential to clarify its clinical relevance and to support more effective management strategies in the early postoperative period of these critically ill patients.

## Conclusion

Although the serum ferritin levels identified in this study remained below the values traditionally associated with severe hyperferritinemia, an association was observed with shorter PICU stays (<7 days) and with PIM 3 scores ≥ 5%. These findings suggest that even moderate elevations in ferritin may reflect systemic inflammatory activation and contribute to early risk stratification in the postoperative management of children with congenital heart defects, particularly in developing countries.

## Data Availability

All data generated in this study are included in this manuscript.

## Acknowledgments

The authors express their sincere appreciation to the medical staff of the Pediatric Intensive Care Unit and the Cardiac Surgery Service at HIMABA for their essential collaboration in facilitating the serum ferritin measurements for the patients enrolled in this study. The authors also extend their gratitude to the patients and their legal guardians for their participation.

